# Relationship between temperature and relative humidity on the initial spread of COVID-19 cases and deaths in Brazil

**DOI:** 10.1101/2021.12.26.21268421

**Authors:** Maria Teresa Anselmo Olinto, Anderson Garcez, Gabriel Brunelli, Flávio Anselmo Olinto, Marcos Fanton, Raquel Canuto

## Abstract

**Introduction:** Climate conditions may have influence on the transmission of COVID-19. Thus, this study aims to evaluate the impact of temperature and relative humidity on COVID-19 cases and deaths during the initial phase of the epidemic in Brazil.

**Methodology:** This is an ecological study based on secondary data. Daily data on new COVID-19 cases and deaths and on climate indicators was collected from February 20th to April 18th, 2020 (n=59 days), for all state capital cities in Brazil and the Federal District (Brasília). The studied climate indicators included mean temperature, temperature amplitude, mean relative humidity, relative humidity amplitude, and percentage of days with mean relative humidity less than or equal to 65%. Correlation and multiple linear regression analysis were performed for all cities and was also stratified by quintiles of the COVID-19 incidence rate.

**Results:** Mean daily temperature was positively correlated with the number of days until the first COVID-19 case was reported. A lower mean relative humidity was correlated with lower number of cases and deaths in Brazil, especially when the relative humidity was less than or equal to 65%. Higher temperatures and humidity amplitudes were correlated with lower COVID-19 mortality. Additionally, after controlling for humidity, cumulative cases of COVID-19 were inversely associated with temperature in cities with mean temperatures less than 25.8 °C.

**Conclusions:** Our findings indicate that variations in temperature and humidity across the Brazilian territory may have influenced the spread of the novel coronavirus during the initial phase of the epidemic in the country.

## Introduction

Coronavirus disease 2019 (COVID-19) is caused by the severe acute respiratory syndrome coronavirus 2 (SARS-CoV-2) [1]. On March 11^th^, 2020, the World Health Organization (WHO) categorized COVID-19 as a pandemic due to the rapid increase of new cases and deaths in different countries [2]. As per the reports on April 1, 2020, there were more than one million confirmed cases and over 50,000 deaths, worldwide [3]. Brazil declared COVID-19 as a public health emergency on February 3^rd^, and the first confirmed case was announced by the Brazilian Ministry of Health on February 25^th^, 2020 [4,5]. The first COVID 19 related death in Brazil was reported on March 17^th^, 2020. Since then, the epidemic has quickly spread countrywide with more than 6,880 cases and 324 deaths per million inhabitants as of April 1, 2020 [6].

Brazil is the largest country in South America, with a population of more than 200 million people [7]. Its territorial expanse is approximately 8.5 million km^2^, and 93% of its landmass lies in the Southern Hemisphere. According to Köppen’s climate classification system, Brazil has three climate zones (tropical, subtropical, and semi-arid) and 12 types of climates [8]. Most of the country’s territory falls in the tropical climate zone (81.4%), except the southern region, which is mainly located in the subtropical climate zone. Brazil exhibits varied climatic conditions, such as dry summers, dry winters, no dry season, hot summers or temperate summers [8].

Climate conditions have been reported to influence the transmission of COVID-19 [9-28]. Upon comparing data on meteorological conditions of 166 countries worldwide, one study concluded that daily new cases and deaths by COVID-19 may be partially suppressed with increase in temperature and humidity [10]. Another study with daily data from several countries also indicated a negative correlation between the average temperature of the country and the number of cases of SARS-CoV-2 infections. An increase in the average daily temperature by one-degree Fahrenheit reduced the number of cases by approximately 6.4 cases/day [14]. Data from 30 provincial capital cities in China revealed that cities with low temperature, mild diurnal temperature range, and low humidity are more likely to favor the transmission [11]. Finally, a systematic review including studies up on April 2020 concluded that cold and dry conditions were potentiating factors on the spread of the virus [29]. However, more recently studies claimed that these associations are not linear [30,31]. Data from England suggests that daily ambient mean temperatures of around 11°C to 13°C pose a higher risk for COVID-19 cases compared to the risk-minimum at 22°C as well as relative humidity showed the highest risk around 61% [31].

Majority of the studies investigating climatic conditions and their impact on COVID-19 have been performed in the Northern Hemisphere [29,32]. Few studies have been conducted in countries located in the Southern Hemisphere, such as Brazil [9,33,34]. One study indicated that when the average temperature was below 25.8°C, a negative linear relationship was observed between temperature and number of COVID-19 confirmed cases. Air humidity was not considered in this analysis [9]. Another case study on the most affected Brazilian cities concluded that high temperatures and intermediate relative humidity might favor the spread of COVID-19 [33].

The relationship between climatic conditions and COVID-19 transmission are not clearly understood and warrants further investigation. There is a lack of studies on COVID 19 in Southern hemisphere, including all Brazilian territory. So, this study aims to explore the impact of temperature and relative humidity on the initial spread of COVID-19 cases and deaths in the initial phase of epidemic in Brazil. It can be interesting to analyze the initial spread period to avoid the multiple factors involved in the COVID-19 pandemic such as community transmission, public health policies, and new variants of SARS-CoV-2.

## Methodology

### Design and study area

This was an ecological study design based on secondary data. The study explored data from all 26 state capital cities of Brazil and the Federal District (Brasília). The cities are located in different regions of Brazil, with different types of climates. The locations of the cities are shown in Figure 1.

### Data collection

Daily data on new cases and deaths by COVID-19, and climate indicators was collected from February 20^th^ to April 18^th^, 2020 (n=59 days), for each state’s capital city and the Federal District (Brasília). Daily cumulative number of newly confirmed cases and deaths by COVID-19 as well as incidence and mortality rates were collected from the official data reported by the Ministry of Health of Brazil (https://covid.saude.gov.br/) [35]. Additionally, data was also collected for the number of days between the start of follow-up (February 20) and the first case reported.

Daily climate data for each city, including mean, maximum, and minimum temperature (°C), and mean, maximum, and minimum relative humidity (%), was collected from the National Institute of Meteorology (http://www.inmet.gov.br/portal/). There was no climate data for the city of Porto Velho in the study period.

The temperature variables included the daily mean temperature and amplitude (maximum value of temperature–minimum value of temperature). The relative humidity variables were daily mean relative humidity and amplitude (maximum value of relative humidity–minimum value of relative humidity). Additionally, two other variables were also proposed based on the potential inactivation of the coronavirus at midrange humidity [36,37]: (1) percentage of days with mean relative humidity less than or equal to 65% in the period (days with relative humidity ≤ 65%/59 × 100); and (2) percentage of days with minimum humidity less than or equal to 65% (days with minimum humidity ≤ 65%/59 × 100).

### Statistical analysis

Descriptive analyses were performed for COVID-19 outcomes and for temperature and relative humidity data, using means and standard errors, amplitude (range), and relative frequencies. Variables were assessed for normality by Shapiro-Wilk’s test and Spearman’s rank correlation coefficient was used to evaluate the correlations between climate variables and COVID-19 outcomes. Correlation analyses was performed for all cities and stratified by quintiles of COVID-19 incidence rate (cities from the medium and lowest quintiles as opposed to cities from the highest quintiles of incidence rate). Sensitivity analysis was conducted to detect potential heterogeneity in the results. The city of São Paulo was excluded from the analysis because it reported the first case of COVID-19 approximately 25 days earlier than the other cities.

Multiple linear regression analysis was also performed to investigate the relationship between daily mean temperature and daily cumulative cases of COVID-19, adjusting for daily mean relative humidity. This analysis was stratified by mean temperature in the period (< 25.8°C vs. ≥ 25.8°C). This cutoff was based on a previous finding, which indicated that each 1°C rise in temperature was associated with a decrease of 4.89% in the number of daily cumulative confirmed cases of COVID-19, only when the average temperature was below 25.8°C [9].

Data analysis was performed using the Statistical Package for Social Sciences version 22.0 (IBM Corp., Armonk, NY, USA). All tests were two-sided, and P < 0.05 was considered statistically significant.

## Results

Summary results for COVID-19 occurrence, temperature, and relative humidity in Brazilian state capitals and the Federal District (Brasília) from February 20th to April 18th, 2020 are shown in Table 1. The COVID-19 cumulative cases are displayed using bubble sizes in Figure 1. The cities in the highest quintile of incidence rates were located in the North and Northeast regions, except for São Paulo, located in the southeast region of Brazil. São Paulo and Rio de Janeiro, the most densely populated cities in Brazil, showed a higher number of cumulative cases, 9,428 and 3,059 cases up till April 18th, respectively. The highest incidence rate was observed in Fortaleza (729 per million inhabitants) with 2,562 cumulative cases. These results were coherent with the time-lapse between the beginning of data collection (5, 15 and 24 days) and the first case reported in São Paulo, Rio de Janeiro, and Fortaleza, respectively. Longer time-lapse (≥ 32 days) until the first case reported resulted in lower incidence rates. The highest mortality rate was observed in São Paulo (45.50 per million inhabitants) (Table 1).

**Table 1.** Descriptive data for the COVID-19 occurrence and daily temperature and relative humidity in Brazilian state capitals and the Federal District (Brasília) from February 20 to April 18, 2020 (n=59 days), according to COVID-19 incidence rate.

| Cities | COVID-19 |  |  |  |  | Temperature (°C) |  | Relative humidity (%) |  |  |  |
| --- | --- | --- | --- | --- | --- | --- | --- | --- | --- | --- | --- |
|  | Incidence rate<sup>a</sup> | Cumulative cases | Mortality rate<sup>a</sup> | Cumulative deaths | Days until first case (lapse) | Mean | Mean amplitude | Mean | Mean amplitude | % days mean≤65<sup>b</sup> | % days min≤65<sup>c</sup> |
| Fortaleza | 729.00 | 2562.00 | 34.80 | 141.00 | 24 | 27.36 | 7.31 | 80.39 | 31.80 | 0.00 | 83.10 |
| São Paulo | 633.70 | 9428.00 | 45.50 | 686.00 | 5 | 20.88 | 9.24 | 70.69 | 41.14 | 27.00 | 91.50 |
| Manaus | 618.50 | 1593.00 | 42.10 | 134.00 | 21 | 27.52 | 7.23 | 78.25 | 34.80 | 0.00 | 86.40 |
| Macapá | 568.20 | 321.00 | 9.90 | 8.00 | 28 | 26.44 | 6.92 | 84.22 | 32.14 | 2.00 | 69.50 |
| Recife | 520.70 | 1255.00 | 37.70 | 106.00 | 21 | 27.58 | 8.79 | 77.29 | 37.27 | 0.00 | 96.60 |
| São Luís | 483.70 | 862.00 | 30.90 | 38.00 | 27 | 26.07 | 5.49 | 90.09 | 26.03 | 0.00 | 27.10 |
| Florianópolis | 413.20 | 221.00 | 6.00 | 3.00 | 22 | 23.35 | 9.06 | 69.18 | 38.05 | 25.00 | 94.90 |
| Rio de Janeiro | 374.90 | 3059.00 | 24.90 | 237.00 | 15 | 24.10 | 5.34 | 80.36 | 19.98 | 0.00 | 33.90 |
| Vitória | 372.80 | 224.00 | 16.60 | 9.00 | 28 | 25.32 | 8.14 | 78.92 | 39.27 | 0.00 | 91.50 |
| Boa Vista | 263.00 | 183.00 | 5.00 | 2.00 | 30 | 29.41 | 11.23 | 56.00 | 41.27 | 90.00 | 100.00 |
| Porto Alegre | 233.20 | 369.00 | 5.40 | 9.00 | 19 | 22.58 | 11.25 | 70.14 | 46.44 | 24.00 | 94.90 |
| Brasília | 226.20 | 762.00 | 5.60 | 24.00 | 16 | 21.25 | 8.78 | 80.20 | 40.34 | 3.00 | 86.40 |
| Natal | 193.40 | 236.00 | 5.70 | 5.00 | 21 | 27.87 | 6.21 | 81.07 | 27.47 | 0.00 | 54.20 |
| Rio Branco | 186.60 | 112.00 | 7.40 | 4.00 | 26 | 25.82 | 8.26 | 85.28 | 41.17 | 0.00 | 72.70 |
| Belém | 180.90 | 449.00 | 10.70 | 22.00 | 26 | 25.87 | 8.30 | 85.79 | 34.81 | 2.00 | 68.20 |
| Salvador | 180.70 | 759.00 | 5.20 | 19.00 | 22 | 27.32 | 5.87 | 80.54 | 23.61 | 0.00 | 39.00 |
| Curitiba | 157.80 | 346.00 | 3.60 | 8.00 | 20 | 19.17 | 11.20 | 68.16 | 54.53 | 36.00 | 94.90 |
| Belo Horizonte | 151.30 | 420.00 | 2.40 | 8.00 | 24 | 20.18 | 9.42 | 84.49 | 37.00 | 0.00 | 76.30 |
| João Pessoa | 142.10 | 148.00 | 17.30 | 17.00 | 27 | 27.81 | 7.31 | 76.90 | 32.90 | 0.00 | 91.50 |
| Cuiabá | 129.00 | 91.00 | 0.00 | 1.00 | 31 | 28.44 | 9.54 | 65.43 | 39.68 | 59.00 | 100.00 |
| Goiânia | 116.10 | 226.00 | 4.60 | 10.00 | 21 | 24.36 | 10.72 | 74.99 | 47.24 | 9.00 | 94.90 |
| Porto Velho<sup>d</sup> | 92.50 | 71.00 | 3.80 | 2.00 | 30 | - | - | - | - | - | - |
| Teresina | 72.80 | 94.00 | 5.80 | 5.00 | 31 | 26.88 | 9.14 | 81.64 | 38.27 | 0.00 | 94.90 |
| Campo Grande | 68.10 | 85.00 | 2.20 | 2.00 | 22 | 25.08 | 11.30 | 62.86 | 43.36 | 68.00 | 96.60 |
| Maceió | 64.80 | 107.00 | 2.90 | 6.00 | 33 | 27.31 | 7.19 | 79.11 | 30.95 | 0.00 | 88.50 |
| Palmas | 56.80 | 23.00 | 3.30 | 1.00 | 32 | 26.73 | 8.54 | 78.08 | 36.80 | 5.00 | 79.20 |
| Aracaju | 56.30 | 52.00 | 6.10 | 4.00 | 32 | 28.41 | 6.84 | 60.94 | 26.34 | 85.00 | 100.00 |

<sup>a</sup> Rate per million inhabitants;
<sup>b</sup> The percentage of days that the average of relative humidity was $\leq 65\%$ ;
<sup>c</sup> The percentage of days that the average minimum of relative humidity was $\leq 65\%$ ;
<sup>d</sup> There is no temperature and humidity data available for Porto Velho, capital of Rondônia State.
The color represents different quintile from high to low quintile of COVID-19 incidence rate.

The highest mean temperature in the period (29.41 °C) was observed in the North region (Boa Vista), and the lowest mean temperature (19.17 °C) was observed in the southern region (Curitiba). The cities from the 5th quintile of the COVID-19 incidence rate showed a mean temperature of approximately 27°C and a mean daily relative humidity greater than 77%, except São Paulo. Curitiba, Porto Alegre, Goiania, Campo Grande, and Boa Vista which showed the largest daily temperature amplitude (around 11°C) as well as the largest daily relative humidity amplitude. None of these capitals showed incidence rates belonging to the highest quintile (Table 1).

Table 2 presents the correlation values for the relationship between COVID-19 occurrence and daily temperature and relative humidity. We observed that mean daily temperature was positively correlated with the number of days until the first COVID-19 case (r = 0.513; p = 0,009) and negatively correlated with mortality (rate and cumulative deaths). Mean daily relative humidity was positively correlated with COVID-19 cumulative cases (r = 0.414; p = 0.040) and deaths (r = 0.408; p = 0,043). The measures of amplitude temperature (r = -0.550; p = 0,004) or humidity (r = -0.473; p = 0.029) were negatively correlated with COVID-19 mortality rates. Humidity amplitude was also negatively correlated with cumulative deaths (r = -0.426; p = 0,034). The percentage of days with a mean humidity less than or equal to 65% were negatively correlated with all COVID-19 measures. After stratification, some correlations were observed, such as humidity amplitude vs. time-lapse until the first case (r = -0.517; p<0.05; negative relation) and others showed more evidence for cities with medium and low COVID-19 incidence rates (Table 2).

**Table 2.**
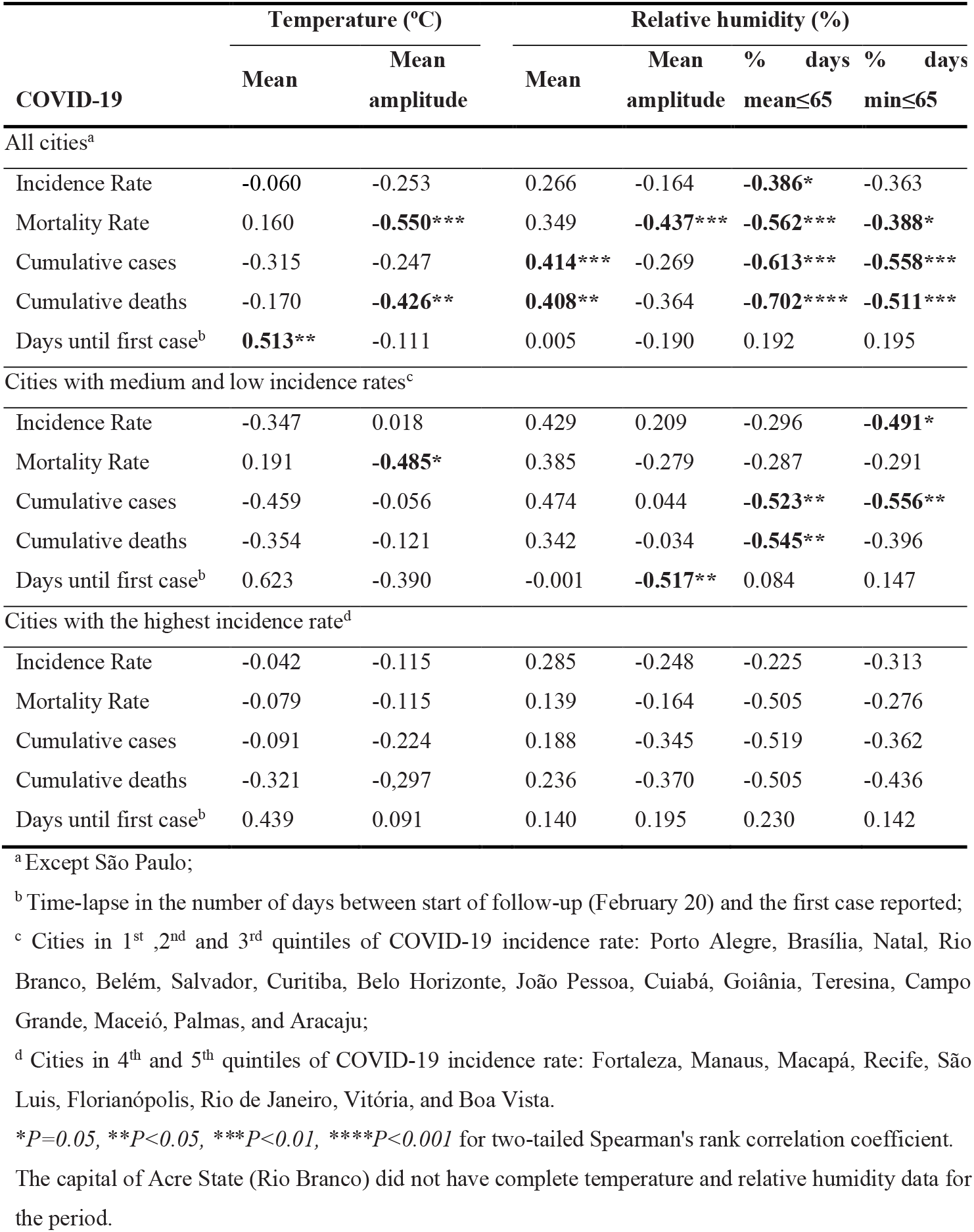
Correlation values for the relationship between COVID-19 occurrence and daily temperature and relative humidity in Brazilian state capitals and the Federal District (Brasília) from February 20 to April 18, 2020.

Linear regression coefficients were obtained for cumulative cases of COVID-19 stratified by mean daily temperature (Table 3). We observed a linear and inverse relationship between COVID-19 cumulative cases and temperature among most cities with mean daily temperatures less than 25.8 °C. This association was evident even after adjusting for relative humidity. The number of COVID-19 cumulative cases decreased by 713.27 for each 1°C increase in temperature in São Paulo. However, this relationship was not observed for cities with a mean daily temperature equal to or greater than 25.8 °C (Table 3).

**Table 3.** Linear regression coefficients (Beta) for cumulative cases of COVID 19 in Brazilian state capitals and the Federal District (Brasília) from February 20 to April 18, 2020, according to mean temperature in the period.

| Cities | $\beta$ | SE | t | p-value | Adjusted $\beta^a$ | SE | t | p-value |
| --- | --- | --- | --- | --- | --- | --- | --- | --- |
| **Cities with mean temperature <25.8°C in the period<sup>b</sup>** |  |  |  |  |  |  |  |  |
| São Paulo | -531.71 | 162.88 | -3.26 | 0.002 | **-713.27** | **154.05** | **-4.63** | **<0.001** |
| Rio de Janeiro | -276.75 | 76.27 | -3.63 | 0.001 | **-345.71** | **78.05** | **-4.43** | **<0.001** |
| Porto Alegre | -31.69 | 4.27 | -7.42 | <0.001 | **-31.65** | **4.78** | **-7.41** | **<0.001** |
| Curitiba | -25.15 | 5.18 | -4.85 | <0.001 | **-26.34** | **5.11** | **-5.15** | **<0.001** |
| Florianópolis | -23.25 | 3.22 | 7.22 | <0.001 | **-22.94** | **3.63** | **-6.31** | **<0.001** |
| Goiânia | -13.17 | 5.73 | -2.28 | 0.026 | **-29.16** | **9.84** | **-2.96** | **0.004** |
| Belo Horizonte | -8.99 | 12.95 | -0.69 | 0.490 | -8.636 | 15.98 | -0.54 | 0.591 |
| Cuiabá | -7.50 | 1.94 | -3.86 | <0.001 | **-14.90** | **2.23** | **-6.66** | **<0.001** |
| Vitória | -5.66 | 4.63 | -1.22 | 0.227 | **-12.53** | **4.77** | **-2.63** | **<0.011** |
| Campo Grande | -5.16 | 1.12 | -4.58 | <0.001 | **-5.92** | **1.12** | **-4.58** | **<0.001** |
| Brasília | -0.74 | 28.18 | -0.026 | 0.979 | -17.32 | 57.95 | -0.29 | 0.766 |
| **Cities with mean temperature $\geq 25.8^\circ\text{C}$ in the period<sup>b</sup>** |  |  |  |  |  |  |  |  |
| Manaus | -140.62 | 41.78 | -3.36 | 0.001 | **-186.16** | **92.58** | **-2.01** | **0.049** |
| Recife | -81.50 | 47.66 | -1.71 | 0.093 | -57.75 | 87.93 | -0.66 | 0.514 |
| Fortaleza | -43.64 | 103.71 | -0.42 | 0.675 | 153.7 | 231.68 | 0.66 | 0.510 |
| Salvador | -42.73 | 28.22 | -1.51 | 0.135 | 35.63 | 45.78 | 0.78 | 0.440 |
| Macapá | -31.47 | 13.26 | -2.37 | 0.021 | -9.94 | 19.89 | -0.50 | 0.619 |
| São Luís | -17.96 | 32.26 | -0.56 | 0.580 | 7.29 | 45.95 | 0.16 | 0.874 |
| Boa vista | -10.98 | 7.23 | -0.19 | 0.134 | -4.90 | 9.31 | -0.52 | 0.602 |
| Natal | -9.41 | 8.90 | -1.05 | 0.295 | 3.26 | 18.64 | 0.17 | 0.862 |
| João Pessoa | -7.74 | 5.40 | -1.43 | 0.157 | -1.96 | 10.95 | -0.18 | 0.858 |
| Aracaju | -2.75 | 1.92 | -1.43 | 0.158 | 2.94 | 3.16 | 0.93 | 0.355 |
| Maceió | -0.46 | 1.43 | -0.33 | 0.746 | **7.31** | **2.57** | **2.84** | **0.007** |
| Belém | 1.73 | 5.90 | 0.29 | 0.770 | 7.13 | 8.12 | 0.88 | 0.385 |
| Teresina | 7.80 | 2.28 | 3.42 | 0.001 | 8.05 | 5.81 | 1.38 | 0.172 |
| Palmas | 0.77 | 0.64 | 1.20 | 0.235 | 0.52 | 1.64 | 0.107 | 0.752 |

<sup>a</sup> Adjusted for daily mean relative humidity in the period;
<sup>b</sup> This cutoff was based on a previous finding (Prata et al., 2020);
The capital of Acre State (Rio Branco) did not have complete temperature and relative humidity data for the period.

## Discussion

The aim of this study was to evaluate possible contributions of temperature and relative humidity variations to the initial spread of the COVID-19 cases and deaths in Brazil. Our results showed that mean daily temperature was positively correlated with the number of days until the first COVID-19 case was reported. A lower mean relative humidity was correlated with lower number of cases and deaths. Higher temperature and humidity amplitudes were correlated with lower COVID-19 mortality. Additionally, after controlling for humidity, cumulative cases of COVID-19 were inversely associated with temperature in cities with mean temperatures less than 25.8 °C.

In this study the initial phase of COVID-19 spread in Brazil was considered to be from February 20^th^ to April 18^th^, 2020. The first day of data collection was defined as the first official day of Carnival, the most popular holiday in the country, which brings together millions of people celebrating in the streets (mainly in São Paulo, Rio de Janeiro, and Salvador) and attracts foreign tourists from Europe and North America. Even with the possibility that SARS-CoV-2 had already been circulating in the country, there was no cancellation of the Carnival. Although the choice of the final date of data collection was arbitrary, it aimed to control for potential effects in the analysis, such as the different public actions to combat COVID-19 in each city and their differences in the healthcare system as well as the possible changes in the season climate.

The Brazilian COVID-19 epidemic started in the three major cities of Brazil: São Paulo, Rio de Janeiro, and Brasília. These cities also have a higher number of international flights in the country. After 54 days of the epidemic, the five Brazilian capital cities with the highest COVID-19 incidence rates were identified in the north (Manaus and Macapá) and northeast regions (Fortaleza and Recife), with the exception of São Paulo [35]. North and Northeast capitals are characterized by a tropical climate with the highest temperature and humidity in Brazil.

Our findings showed that a lower mean relative humidity was correlated with a lower number of cases and deaths. Previous studies have also explored this association [10,11,16,20,24,25,38-40]. However, some studies have shown an inverse relationship between relative humidity and new cases and deaths [10,16,25,41]. Analyzing the amplitudes of relative humidity, a study demonstrated that high epidemic transmission of COVID-19 is associated with the humidity range of 60% - 90% [31,38]. This study included information from January 20^th^ and March 11^th^ for 430 cities and districts in China, 21 cities/provinces in Italy, 21 cities/provinces in Japan, and 51 other countries around the world. Another study with Chinese data demonstrated a humidity range of 50% - 80% is conducive to the survival of the coronavirus [39], whereas data from England demonstrated a highest risk around 61% of relative humidity [31]. In our analyses, Brazilian cities with mean humidity less than or equal to 65% had lower mortality rates, cumulative cases, and cumulative deaths. Additionally, the correlation between relative humidity and COVID-19 occurrence was more evident in cities with higher incidence rates up till April 18^th^, demonstrating that this relationship can be more important at the beginning of the epidemic process.

Furthermore, our results revealed that higher humidity amplitudes were correlated with lower COVID-19 mortality rates. Restricting analysis only for capitals with medium and low incidence rates showed a negative correlation between humidity amplitude and time-lapse until the first case. Therefore, it exhibits a complex relation which needs to be further investigated. A laboratory study, which investigated how virus survival is affected by air temperature and relative humidity, has revealed a potential inactivation of the coronavirus at midrange relative humidity (∼50%), and a greater protective effect of the virus at low (20%) and high (80%) relative humidity [36]. Thus, the higher humidity amplitude has a higher probability of including these extreme values of relative humidity, leading to a greater potential for virus transmissibility inactivation.

Temperature also has an impact on the transmission of COVID-19. In our study, higher temperature showed a positive association with a larger time-lapse between the beginning of data collection and the first COVID-19 case reported. Studies using data from only northern hemisphere cities have shown that higher temperatures could reduce the spread of the virus [11,25,42]. Another study exploring data from 166 different countries in all hemispheres demonstrated a linear negative correlation between temperature and COVID-19 cases [10]. This finding could be biased by the fact that the pandemic had started in the coldest seasons of the year for the northern hemisphere. Consequently, these countries had a greater number of cases compared to southern hemisphere countries. Similarly, Wang’s study, which included data from China and the United States, showed that every 1 °C increase in average temperature led to an increase of 0.83 cumulative number of cases at lower temperature. However, with higher temperatures in summer, this effect will probably decrease [16].

Previous data from a tropical climate country (Brazil) revealed a negative linear relationship between temperature and COVID-19 cases, only when the mean temperature was below 25.8°C [9]. This suggests a possible nonlinear dose-response relationship between the two variables, indicating that the effect of temperature on mitigating the spread of the COVID-19 is limited to colder temperatures. Our results corroborate these findings because cumulative cases of COVID-19 were inversely associated with temperature only in cities with mean temperatures less than 25.8 °C.

In summer, higher temperature amplitudes can be observed in subtropical climate cities in the South and Midwest regions of Brazil. In this study, cities with high amplitude were negatively correlated with COVID-19 deaths (rate and cumulative). Previous studies indicated that the diurnal temperature range could affect COVID-19 transmission [11,25]. A positive association with COVID-19 daily death counts was observed for diurnal temperature range in a study conducted in Wuhan, China [25]. However, another study in China revealed that a 1 °C increase in diurnal temperature range resulted in decline of daily confirmed case counts [11]. This suggests that an environment with a small daily amplitude or constant temperature could be more advantageous for virus survival. However, these effects were analyzed in cold climates and without control for relative humidity.

In this study, cumulative cases of COVID-19 were inversely associated with temperature in cities with mean temperature less than 25.8 °C, after controlling the association with humidity. The number of COVID-19 cumulative cases decreased by 713.27 for each 1 °C increase in temperature in São Paulo. Our results indicate a significant interaction between temperature and humidity. In accordance with this finding, a study reported a significant interaction between daily average temperature and relative humidity for the COVID-19 incidence in Hubei province, Mainland China [20]. Temperature and humidity showed negative associations with COVID-19. Every 1 °C increase in the daily average temperature led to a decrease in the daily confirmed cases by 36% to 57% when relative humidity was in the range of 67% to 85.5%. Every 1% increase in relative humidity led to a decrease in the daily confirmed cases by 11% to 22% when the daily average temperature was in the range of 5.04 to 8.2 °C [20]. Another study also indicated that there is some influence of the interaction of average temperature and average relative humidity on the incidence of COVID-19 in India. However, this finding was not consistent throughout the study area [28]. Although the mechanism of this interaction remains unclear, regions with low temperature and humidity are more affected by the virus. Meteorological factors can create ideal conditions for virus attachment, replication, and transmission as well as their survival in the environment.

This is the first study to show how the combination of temperature and relative humidity can influence the COVID-19 initial spread and mortality in all Brazilian territories. We included several meteorological values for temperature and relative humidity in this study. Daily meteorological data was used to appropriately compare the effect of temperature and humidity on the COVID-19 cases and deaths. Additionally, we evaluated this relationship in the initial period of the COVID-19 pandemic. In order to dismiss possible effects of the underreporting and city population density on the results, we adopted different outcome measures, including cases and deaths. The results were consistent with all the measures. Nevertheless, some methodological considerations should be addressed. First, as an ecological study, it has limitations in establishing causal relationships, as the association observed on an aggregate level does not necessarily represent the association that exists at an individual level. However, ecologic analysis may be more appropriate than studies using individual data when investigating the determinants of transmission of infectious diseases with complex and nonlinear infection spread [43]. Second, we did not include ultraviolet radiation in the analyses, although recent studies provide that ultraviolet radiation is associated with incidence rates of COVID 19 [44,45]. Third, it was not our goal to assess other determinants for the spread and mortality of COVID-19, such as social distancing strategies, health system structure, medical resources, people’s endurance, and personal hygiene. These aspects should be examined in future studies. Fourth, we sought to explore data for all Brazilian state capitals and the Federal District from February 20^th^ to April 18^th^, 2020. However, we could not obtain the meteorological data for Porto Velho, capital of Rondônia State. Finally, further investigations in the countryside cities of Brazil should be explored if a novel epidemic spread occurs. Replication of this investigation might lead to more conclusive results using data from the end of winter, especially for cities located in the southern region of the country.

## Conclusion

Variations in temperature and humidity across the Brazilian territory may have influenced the spread of the novel coronavirus during the initial phase of the epidemic in the country. Our study revealed that lower relative humidity was correlated with a lower number of cases and deaths in Brazil, especially when the relative humidity was less than or equal to 65%. Higher temperature and humidity amplitudes were also correlated with lower COVID-19 cases and deaths. In addition, after controlling for humidity, cumulative cases of COVID-19 were inversely associated with temperature in cities with mean temperatures less than 25.8 °C. Although large-scale implementation of public health control measures plays a considerable role in the reduction of COVID-19 cases and deaths, our results indicate an independent contribution of climate conditions on the initial spread of the COVID-19.

## Data Availability

All data produced in the present study are available upon reasonable request to the authors

## Acknowledgements

M.T.A.O. received research productivity grants from the Brazilian Council for Scientific and Technological Development - CNPq (process n. 307175/2017-0). A.G. received a post-doctoral fellowship from CNPq (process n. 161302/2019-0).

## Supplementary files legends

**Figure 1.** COVID-19 cumulative cases (bubble size) in Brazilian state capitals and the Federal District (Brasília) from February 20 to April 18, 2020.

